# Exposure to air pollution and COVID-19 mortality in the United States: A nationwide cross-sectional study

**DOI:** 10.1101/2020.04.05.20054502

**Authors:** Xiao Wu, Rachel C Nethery, M Benjamin Sabath, Danielle Braun, Francesca Dominici

## Abstract

**Objectives:** United States government scientists estimate that COVID-19 may kill tens of thousands of Americans. Many of the pre-existing conditions that increase the risk of death in those with COVID-19 are the same diseases that are affected by long-term exposure to air pollution. We investigated whether long-term average exposure to fine particulate matter (PM^2.5^) is associated with an increased risk of COVID-19 death in the United States.

**Design:** A nationwide, cross-sectional study using county-level data.

**Data sources:** COVID-19 death counts were collected for more than 3,000 counties in the United States (representing 98% of the population) up to April 22, 2020 from Johns Hopkins University, Center for Systems Science and Engineering Coronavirus Resource Center.

**Main outcome measures:** We fit negative binomial mixed models using county-level COVID-19 deaths as the outcome and county-level long-term average of PM^2.5^ as the exposure. In the main analysis, we adjusted by 20 potential confounding factors including population size, age distribution, population density, time since the beginning of the outbreak, time since state’s issuance of stay-at-home order, hospital beds, number of individuals tested, weather, and socioeconomic and behavioral variables such as obesity and smoking. We included a random intercept by state to account for potential correlation in counties within the same state. We conducted more than 68 additional sensitivity analyses.

**Results:** We found that an increase of only 1 *μ*g/m^3^ in PM^2.5^ is associated with an 8% increase in the COVID-19 death rate (95% confidence interval [CI]: 2%, 15%). The results were statistically significant and robust to secondary and sensitivity analyses.

**Conclusions:** A small increase in long-term exposure to PM^2.5^ leads to a large increase in the COVID-19 death rate. Despite the inherent limitations of the ecological study design, our results underscore the importance of continuing to enforce existing air pollution regulations to protect human health both during and after the COVID-19 crisis. The data and code are publicly available so our analyses can be updated routinely.

**Summary Box:** *What is already known on this topic:*

1. Long-term exposure to PM^2.5^ is linked to many of the comorbidities that have been associated with poor prognosis and death in COVID-19 patients, including cardiovascular and lung disease.
2. PM^2.5^ exposure is associated with increased risk of severe outcomes in patients with certain infectious respiratory diseases, including influenza, pneumonia, and SARS.
3. Air pollution exposure is known to cause inflammation and cellular damage, and evidence suggests that it may suppress early immune response to infection.

*What this study adds:*

1. This is the first nationwide study of the relationship between historical exposure to air pollution exposure and COVID-19 death rate, relying on data from more than 3,000 counties in the United States. The results suggest that long-term exposure to PM^2.5^ is associated with higher COVID-19 mortality rates, after adjustment for a wide range of socioeconomic, demographic, weather, behavioral, epidemic stage, and healthcare-related confounders.
2. This study relies entirely on publicly available data and fully reproducible, public code to facilitate continued investigation of these relationships by the broader scientific community as the COVID-19 outbreak evolves and more data become available. A small increase in long-term PM^2.5^ exposure was associated with a substantial increase in the county’s COVID-19 mortality rate up to April 22, 2020.

## Introduction

The scale of the COVID-19 public health emergency is unmatched in our lifetime and will have grave social and economic consequences. The suddenness and global scope of this pandemic has raised urgent questions that require coordinated investigation in order to slow the disease’s devastation. A critically important public health objective is to identify key modifiable environmental factors that may contribute to the severity of the health outcomes (e.g., ICU hospitalization and death) among individuals with COVID-19. Data from China and Italy show that a majority of COVID-19 deaths occurred in adults aged ≥60 years^1^ and in persons with serious underlying health conditions.^2-4^ Early age-stratified COVID-19 death rates in the United States, reported by the Centers for Disease Control and Prevention (CDC),^5^ also suggest that persons aged ≥65 are at highest risk. Additional factors associated with severe disease include male sex and the presence of comorbidities including hypertension, obesity, diabetes mellitus, cardiovascular disease, and chronic lung disease.^6 7^ Severe COVID-19 infection is characterized by a high inflammatory burden, and it can cause viral pneumonia with additional extrapulmonary manifestations and complications including acute respiratory distress syndrome (ARDS),^8-13^ which has a mortality rate ranging from 27% to 45%.^14^ Studies have also documented high rates of heart damage,^11 15^ cardiac arrhythmias,^12^ and blood clots^16^ in COVID-19 patients. Patients with severe disease can suffer respiratory failure and failure of other vital systems, leading to death.

Although the epidemiology of COVID-19 is evolving, there is a large overlap between causes of death in COVID-19 patients and the conditions caused and/or exacerbated by long-term exposure to fine particulate matter (PM^2.5^). PM^2.5^ contains microscopic solids or liquid droplets small enough that they can be inhaled and cause serious health problems. The Global Burden of Disease Study identified air pollution as a risk factor for total and cardiovascular disease mortality, and it is believed to have contributed to nearly 5 million premature deaths worldwide in 2017 alone.^17^ On Thursday, March 26, 2020 the US EPA announced a sweeping relaxation of environmental rules in response to the coronavirus pandemic, allowing power plants, factories and other facilities to determine for themselves if they are able to meet legal requirements on reporting air and water pollution. The association between PM^2.5^ and health, including both infectious and chronic respiratory diseases, cardiovascular diseases, neurocognitive disease, and pregnancy outcomes in the United States and worldwide is well established.^18-24^ A recent study by our group also documented a statistically significant association between long-term exposures to PM^2.5^ and ozone and risk of ARDS among older adults in the United States.^25^ Numerous scientific studies reviewed by the United States Environmental Protection Agency (US EPA) have linked PM^2.5^ to a variety of health concerns including premature death in people with heart or lung disease, non-fatal heart attacks, irregular heartbeats, aggravated asthma, decreased lung function, and increased respiratory symptoms such as inflammation, airway irritations, coughing, or difficulty breathing.^26^

We hypothesize that because long-term exposure to PM^2.5^ adversely affects the respiratory and cardiovascular systems and increases mortality risk,^27-29^ it also exacerbates the severity of COVID-19 infection symptoms and worsens the prognosis of COVID-19 patients. In this study, we quantified the impact of long-term PM^2.5^ exposure on COVID-19 mortality rates in United States counties. Our study includes 3,087 counties in the United States, covering 98% of the population. We leveraged our previous efforts that focused on estimating the long-term effects of PM^2.5^ on mortality among 60 million United States’ Medicare enrollees.^20 30 31^ We used a well-tested research data platform that gathers, harmonizes, and links nationwide air pollution data, census data, and other potential confounding variables with health outcome data. We augmented this platform with newly collected COVID-19 data from authoritative data sources.^32^ All data sources used in these analyses, along with fully reproducible code, are publicly available to facilitate continued investigation of these relationships as the COVID-19 outbreak evolves and more data become available.

## Methods

Table 1 summarizes our data sources and their provenance, including links where the raw data can be extracted directly.

**Table 1:** Publicly available data sources used in the analysis

|  | Source | Data |
| --- | --- | --- |
| Outcome: COVID-19 Deaths | Johns Hopkins University the Center for Systems Science and Engineering (JHU-CSSE) Coronavirus Resource Center (https://coronavirus.jhu.edu/) | County-level COVID-19 death count up to and including April 22, 2020 |
| Exposure: PM<sub>2.5</sub> concentrations | Atmospheric Composition Analysis Group (https://sites.wustl.edu/acag/) | 0.01° × 0.01° grid resolution PM<sub>2.5</sub> prediction, averaged across the period 2000–2016 and averaged across grid cells in each county |
| Confounders for main analysis | US Census/American Community Survey (https://www.census.gov/programs-surveys/acs/data.html) | County-level socioeconomic and demographic variables for 2012–2016 |
|  | Robert Wood Johnson Foundation County Health Rankings (https://www.countyhealthrankings.org/) | County-level behavioral risk factor variables for 2020 |
|  | JHU-CSSE Coronavirus Resource Center | Time since first reported COVID-19 case |
|  | Raifman et al, Boston University School of Public Health, COVID-19 United States state policy database (www.tinyurl.com/statepolicies) | Time since issuance of stay-at-home order |
|  | Homeland Infrastructure Foundation-Level Data (HIFLD) (https://hifld-geoplatform.opendata.arcgis.com/datasets/hospitals) | County-level number of hospital beds in 2019 |
|  | Gridmet via Google Earth engine (https://developers.google.com/earth-engine/datasets/catalog/IDAHO_EPSCOR_GRIDMET) | 4 km × 4 km temperature and relative humidity predictions, summer and winter averaged across the period 2000–2016 and averaged across grid cells in each county |
| Additional confounders for secondary analyses | The COVID tracking project (https://covidtracking.com/) | State level number of COVID-19 tests performed up to and including April 22, 2020 |
|  | Carnegie Mellon University<br>Delphi Research Center<br>(https://covid-survey.dataforgood.fb.com/) | Estimated percentage of people with COVID-19 symptoms, based on survey data |

### COVID-19 deaths

We obtained COVID-19 death counts for each county in the United States from Johns Hopkins University, Center for Systems Science and Engineering Coronavirus Resource Center.^32^ This source provides the most comprehensive county-level COVID-19 data to date reported by the CDC and state health departments, including the number of new and cumulative deaths and confirmed cases reported in each county across the United States, updated daily. We collected the cumulative number of deaths for each county up to and including April 22, 2020. County-level COVID-19 mortality rates were defined for our analyses as the ratio of COVID-19 deaths to county level population size. While individual-level data would have allowed a more rigorous statistical analyses, individual-level data on COVID-19 death is currently not available.

### Exposure to air pollution

We calculated county-level long-term exposure to PM^2.5^ (averaged from 2000 to 2016) from an established exposure prediction model.^33^ The PM^2.5^ exposure levels were estimated monthly at 0.01° × 0.01° grid resolution across the entire continental United States by combining satellite, modeled, and monitored PM^2.5^ data in a geographically weighted regression. These estimates have been extensively cross-validated.^33^ We aggregated these levels spatially by averaging the values for all grid points within a zip code and then averaging across zip codes within a county. We obtained temporally averaged PM^2.5^ values (2000−2016) at the county level by averaging estimated PM^2.5^ values within a given county. We computed the average 2016 PM^2.5^ exposure analogously for each county to use in sensitivity analyses.

### Potential confounders

In the main analysis, we considered the following 19 county-level variables and one state-level variable as potential confounders (see also Table 2): days since first COVID-19 case reported (a proxy for epidemic stage), population density, percent of population ≥65 years of age, percent of the population 45-64 years of age, percent of the population 15-44 years of age, percent living in poverty, median household income, percent black, percent Hispanic, percent of the adult population with less than a high school education, median house value, percent of owner-occupied housing, percent obese, percent current smokers, number of hospital beds per unit population, and average daily temperature and relative humidity for summer (June-September) and winter (December-February) for each county, and days since issuance of stay-at-home order for each state. Note that publicly available daily COVID-19 case counts at the county level were only available starting March 22, 2020, so that the measure of days since first COVID-19 case reported was truncated by this date. Additional detail on the creation of all variables used in the analysis is available in the Supplementary Materials.

**Table 2:** Characteristics of the study cohort up to and including April 22, 2020, mean (standard deviation)

|  | Total<br>3,087 counties | PM<sub>2.5</sub> <8 µg/m<sup>3</sup><br>1,217 counties | PM<sub>2.5</sub> ≥8 µg/m<sup>3</sup><br>1,870 counties |
| --- | --- | --- | --- |
| COVID-19 death rate (per 100,000) | 3.4 (10.6) | 1.6 (5.7) | 4.7 (12.7) |
| Average PM<sub>2.5</sub> (µg/m<sup>3</sup>) | 8.4 (2.5) | 5.7 (1.4) | 10.1 (1.2) |
| Rate of hospital beds (per 100,000) | 242 (391.9) | 300 (515.2) | 204.2 (278) |
| Days since first case | 23.6 (10.7) | 19 (12.6) | 26.5 (7.9) |
| Days since stay-at-home order | 18.3 (12.4) | 16.7 (13.6) | 19.2 (11.4) |
| % Smokers | 17.4 (3.5) | 15.8 (3.1) | 18.5 (3.4) |
| % Obese | 32.9 (5.4) | 31.2 (5.1) | 34 (5.3) |
| % In poverty | 10.5 (5.7) | 9.7 (5.7) | 11.1 (5.6) |
| % Less than high school education | 21.2 (10.4) | 16.5 (8.7) | 24.2 (10.3) |
| % Owner-occupied housing | 74.2 (8.8) | 76 (7.7) | 73.1 (9.3) |
| % Hispanic | 7.6 (12.3) | 9.7 (13.7) | 6.3 (11.1) |
| % Black | 8.2 (14.2) | 1 (1.8) | 12.9 (16.5) |
| % ≥65 years of age | 16 (4.1) | 17.4 (4.5) | 15 (3.4) |
| % 45-64 years of age | 26.4 (3) | 26.9 (3.8) | 26.1 (2.4) |
| % 15-44 years of age | 37.6 (6.5) | 35.2 (8.2) | 39.2 (4.5) |
| Population density (person/sq. mi.) | 406.7 (1732.6) | 132.6 (430.7) | 585.1 (2180.6) |
| Median household income (\$1,000) | 49 (13.1) | 50.5 (10.9) | 48 (14.3) |
| Median house value (\$1,000) | 136 (89.4) | 140.4 (87.3) | 133.1 (90.6) |
| Average summer temperature (°F) | 86 (5.7) | 83.7 (6.7) | 87.4 (4.4) |
| Average winter temperature (°F) | 45.1 (11.9) | 39.4 (11.5) | 48.7 (10.7) |
| Average summer relative humidity (%) | 89 (9.6) | 83.2 (11.5) | 92.8 (5.5) |
| Average winter relative humidity (%) | 87.5 (4.8) | 87.9 (5.6) | 87.2 (4.1) |

### Statistical methods

We fit a negative binomial mixed model^34-36^ using COVID-19 deaths as the outcome and PM^2.5^ as the exposure of interest to estimate the association between COVID-19 mortality rate and long-term PM^2.5^ exposure, adjusted by covariates. The model included a population size offset and was adjusted for all the potential confounders listed above. We also included a random intercept by state to account for potential correlation in counties within the same state, due to similar socio-cultural, behavioral, and healthcare system features and similar COVID-19 response and testing policies. Additional modeling details are provided in the Supplementary Materials. We report mortality rate ratios (MRR), i.e., exponentiated parameter estimates from the negative binomial model, and 95% CI. The MRR for PM^2.5^ can be interpreted as the relative increase in the COVID-19 mortality rate associated with a 1 *μ*g/m^3^ increase in long-term average PM^2.5^ exposure. We carried out all analyses in R statistical software and performed model fitting using the lme4 package.^37 38^

### Quantifying unmeasured confounding bias

Because this study is observational and the contributing factors to COVID-19 spread and severity remain largely unknown at this early stage of the pandemic, unmeasured confounding is a concern in our analyses. The E-value is a commonly used metric to evaluate the potential impact of unmeasured confounding on results from an observational study.^39^ For a pre-specified exposure variable of interest (long-term exposure to PM^2.5^), the E-value quantifies the minimum strength of association that an unmeasured confounder must have, with both the outcome (COVID-19 mortality rate) and exposure (long-term exposure to PM^2.5^) conditional to all of the potential confounders included in the regression model, to explain away the estimated exposure-outcome relationship. We report the E-value for the MRR estimate for PM^2.5^ under the main model with 20 potential confounders.

### Secondary analyses

In addition to the main analysis, we conducted six secondary analyses to assess the robustness of our results to the confounder set used, outliers, and the model form specification.

First, because the New York metropolitan area has experienced the most severe COVID-19 outbreak in the United States to date, we anticipated that it would strongly influence our analysis. As a result, we repeated the analysis excluding the counties comprising the New York metropolitan area, as defined by the Census Bureau.

Second, although in our main analysis we adjusted for days since first COVID-19 case reported to capture the size of an outbreak in a given county, this measure is imprecise. To further investigate the potential for residual confounding bias (i.e., if counties with high PM^2.5^ exposure also tend to have large outbreaks relative to the population size, then their death rates per unit population could appear differentially elevated, inducing a spurious correlation with PM^2.5^), we also conducted analyses excluding counties with fewer than 10 confirmed COVID-19 cases.

Third, we omitted an anticipated strong confounder, days since first COVID-19 case reported, from the model. Fourth, we additionally adjusted our models for the number of tests performed at the state level (see Table 1 for data source) to evaluate how state-level differences in testing policies might impact our results. Fifth, we additionally adjusted our models for county-level estimated percentage of people with COVID-19 symptoms (see Table 1 for data source) to evaluate how the size of the outbreak in each county might impacts our results. Sixth, we introduced PM^2.5^ into our models as a categorical variable, categorized at the empirical quintiles, to assess the sensitivity of our results to the assumption of a linear effect of PM^2.5^ on COVID-19 mortality rates.

### Sensitivity analyses

We conducted 68 sensitivity analyses to assess the robustness of our results to data and modeling choices. First, we repeated all the analyses using alternative methods to estimate exposure to PM^2.5 31^ Second, we fit the models, modifying the adjustment for confounders, such as using a log transformation or categorized versions of some of the covariates. Third, because our study relies on observational data, our results could be sensitive to modeling choices (e.g., distributional assumptions or assumptions of linearity). We evaluated sensitivity to such choices by considering alternative model specifications and by fitting models stratified by county urban-rural status. Additional detail about the sensitivity analyses and the results are provided in the Supplementary Materials.

## Results

Our study utilized data from 3,087 counties, of which 1,799 (58.3%) had reported zero COVID-19 deaths at the time of this analysis. Table 2 describes the data used in our analyses. All COVID-19 death counts (a total of 45,817 deaths) are cumulative up to April 22, 2020. Figure 1 illustrates the spatial variation of long-term average exposure to PM^2.5^ and COVID-19 death rates (per 1 million population) by county. Visual inspection suggests higher COVID-19 death rates in the Mid-Atlantic, upper Midwest, and Gulf Coast regions. These spatial patterns in COVID-19 death rates generally mimic patterns in both high population density and high PM^2.5^ exposure areas. In the Supplementary Materials, we provide additional data diagnostics that justify the use of the negative binomial model for our analyses.

**Fig 1:**
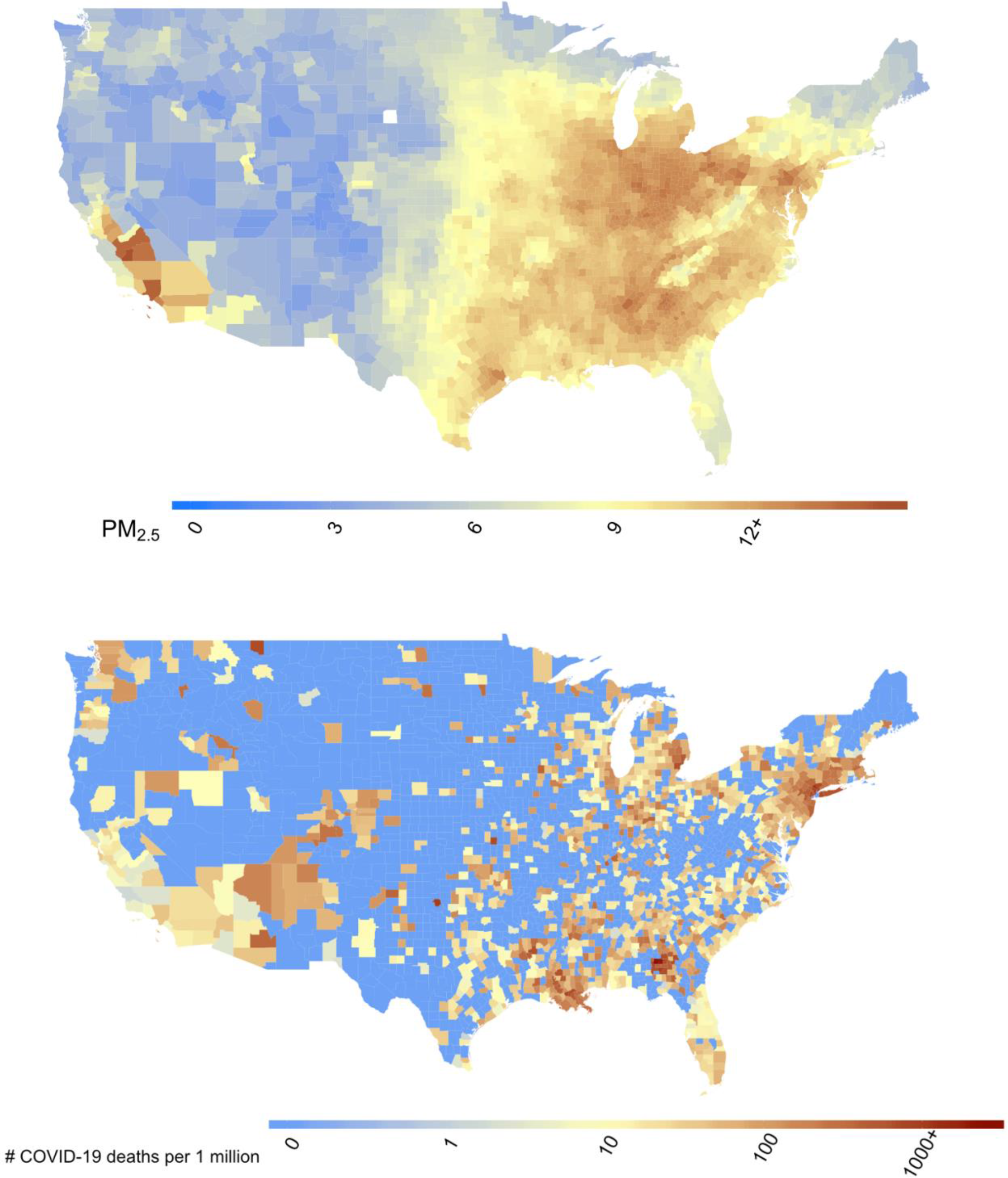
Maps show **(a)** county-level 17-year long-term average of PM^2.5^ concentrations (2000− 2016) in the United States in *μ*g/m^3^, and **(b)** county-level number of COVID-19 deaths per 1 million population in the United States up to and including April 22, 2020.

In Table 3, we report the estimated regression coefficients for each of the covariates included in our main analysis, including PM^2.5.^ We found that the estimated MRR for PM^2.5^ is 1.08 (1.02, 1.15). That is, we found that an increase of only 1 *μ*g/m^3^ in long-term average PM^2.5^ is associated with a statistically significant 8% increase in the COVID-19 death rate. Importantly, we also found that population density, days since first COVID-19 case reported, rate of hospital beds, median household income, percent with less than a high school education, and percent Black are important predictors of COVID-19 death rate. Our results are consistent with previously reported findings that Black Americans are at higher risk of COVID-19 mortality than other groups,^40^ we found a 45% (32%, 60%) increase in COVID-19 mortality rate associated with a 1-standard deviation (per 14.2%) increase in percent Black residents.

**Table 3:** Mortality rate ratios (MRR), 95% confidence intervals (CI), and P-values for all variables in the main analysis.

|  | MRR | 95% CI | P-value |
| --- | --- | --- | --- |
| PM<sub>2.5</sub> ($\mu\text{g}/\text{m}^3$) | 1.08 | (1.02, 1.15) | 0.01 |
| Population density (Q2) | 0.86 | (0.60, 1.23) | 0.40 |
| Population density (Q3) | 0.58 | (0.40, 0.82) | 0.00 |
| Population density (Q4) | 0.47 | (0.33, 0.68) | 0.00 |
| Population density (Q5) | 0.52 | (0.35, 0.77) | 0.00 |
| % Poverty | 1.02 | (0.93, 1.13) | 0.65 |
| log(Median house value) | 1.17 | (0.99, 1.39) | 0.06 |
| log(Median household income) | 1.28 | (1.09, 1.51) | 0.00 |
| % Owner-occupied housing | 1.12 | (1.02, 1.23) | 0.18 |
| % Less than high school education | 1.36 | (1.21, 1.52) | 0.00 |
| % Black | 1.45 | (1.32, 1.60) | 0.00 |
| % Hispanic | 1.00 | (0.89, 1.12) | 0.99 |
| % $\geq 65$ years of age | 1.15 | (0.99, 1.33) | 0.07 |
| % 15-44 years of age | 0.93 | (0.74, 1.17) | 0.54 |
| % 45-64 years of age | 0.96 | (0.83, 1.12) | 0.62 |
| Days since stay-at-home order | 1.28 | (0.97, 1.70) | 0.08 |
| Days since first case | 2.96 | (2.50, 3.51) | 0.00 |
| Rate of hospital beds | 1.12 | (1.02, 1.23) | 0.01 |
| % Obese | 0.94 | (0.86, 1.02) | 0.14 |
| % Smokers | 1.08 | (0.92, 1.26) | 0.36 |
| Average summer temperature ($^{\circ}\text{F}$) | 0.96 | (0.79, 1.16) | 0.68 |
| Average winter temperature ($^{\circ}\text{F}$) | 1.18 | (0.90, 1.53) | 0.22 |
| Average summer relative humidity (%) | 0.84 | (0.71, 1.01) | 0.07 |
| Average winter relative humidity (%) | 1.00 | (0.89, 1.13) | 0.99 |

For our main analysis, the E-value for the estimated MRR for PM^2.5^ was 1.37. That is, in order for an unmeasured confounder to fully account for the estimated effect of PM^2.5^ on the COVID-19 mortality rate, it would have to be associated with both long-term PM^2.5^ exposure and COVID-19 mortality by a risk ratio of at least 1.37-fold each, through pathways independent of all covariates already included in the model. If we were to include such a confounder in our models, along with all other confounders considered, the estimated MRR for PM^2.5^ mortality would become 1 (the null value). To get a sense of the magnitude of the required confounding effect, we also computed the E-value for some of our key measured confounders for comparison. The E-values for days since first COVID-19 case reported (1.16), the weather variables (1.02), number of hospital beds (1.04) and the behavioral risk factors (1.02) were significantly smaller than the reported E-values for the required unmeasured confounder. This suggests that any unmeasured confounder would need to have a confounding effect substantially larger than any of our observed confounders in order to explain away the relationship between PM^2.5^ and COVID-19 mortality rate.

In Figure 2, we report the MRR and 95% CI for PM^2.5^ from all secondary analyses. In these analyses, we separately (a) omitted New York metropolitan area; (b) excluded counties with fewer than 10 confirmed COVID-19 cases; (c) omitted time since first reported COVID-19 case from the model; (d) additionally adjusted the model for number of tests performed; (e) additionally adjusted the model for estimated percentage of people with COVID-19 symptoms; and (f) treated PM^2.5^ as a categorical variable. The results of these analyses were consistent with the main analysis. For the analysis of the PM^2.5^ categorized into quintiles, the MRR for the k^th^ can be interpreted as the increase in COVID-19 mortality rate associated with a change from the first quintile to the k^th^ quintile in long-term PM^2.5^ exposure. The MRR estimates from this model monotonically increased as PM^2.5^ increased, supporting the assumption of a linear relationship between PM^2.5^ and COVID-19 mortality rates. The results of all sensitivity analyses are provided in the Supplementary Materials.

**Fig 2:**
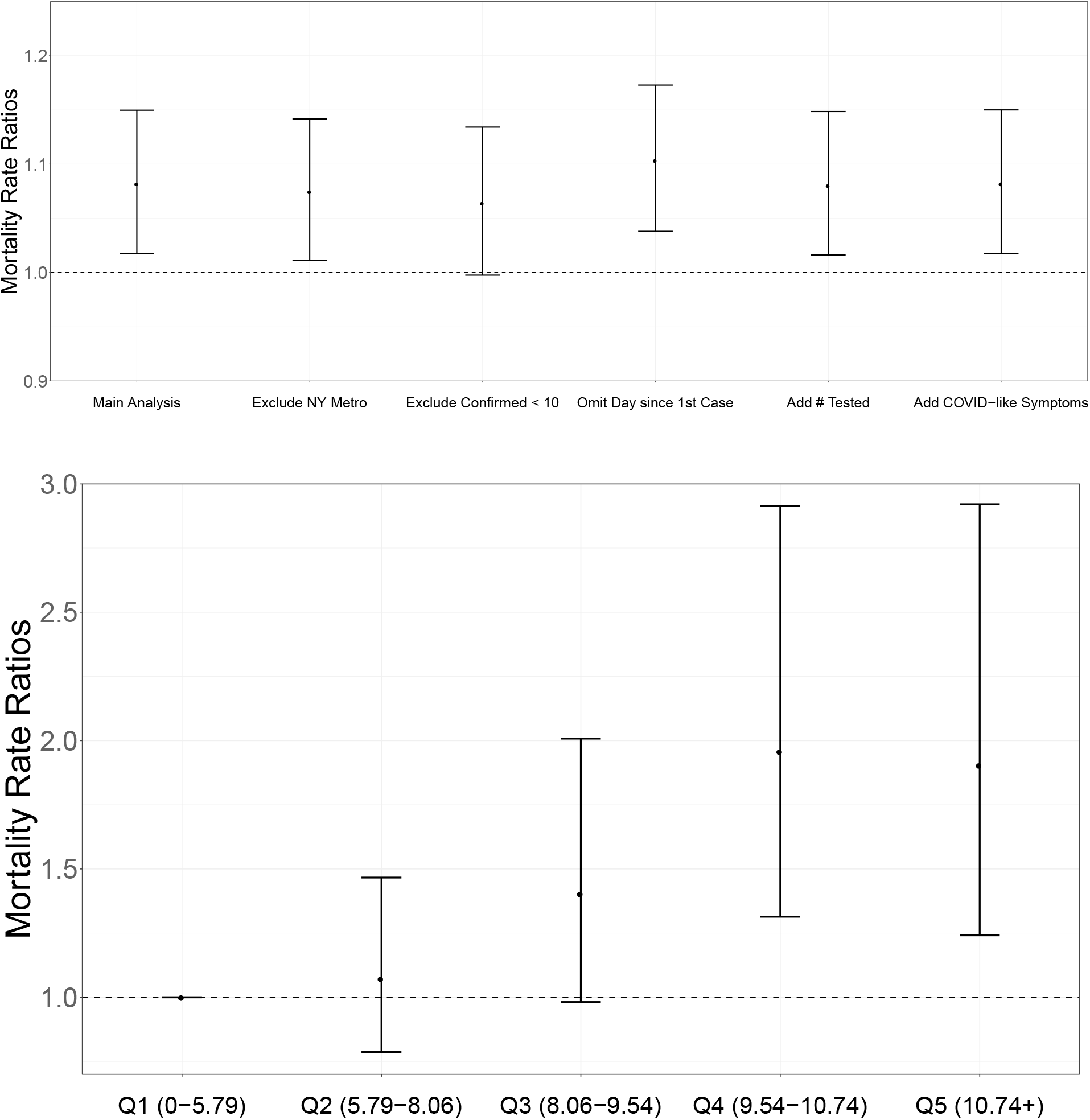
Mortality Risk Ratios (MRR) and 95% confidence intervals. **Upper panel**, MRR can be interpreted as percentage increase in the COVID-19 death rate associated with a 1 *μ*g/m^3^ increase in long-term average PM^2.5^ exposure. The MRR from the main analysis was adjusted for 20 potential confounders. In addition to the main analysis, results are shown for secondary analyses (a) excluding the counties in New York metropolitan area, (b) excluding counties with fewer than 10 confirmed COVID-19 cases, (c) omitting time since first reported COVID-19 case from the model, (d) adding state-level number of tests performed to the model, (e) adding county-level estimated percentage of people with COVID-19 symptoms to the model, and (f) using PM^2.5^ exposure categorized at quintiles. All COVID-19 death counts are cumulative counts up to and including April 22, 2020. **Lower panel**, MRR can be interpreted as the percentage increase in the COVID-19 death rate associated with each empirical quintile of long-term average PM^2.5^ exposure compared to the baseline quintile (Q1).

## Discussion

This is the first nationwide study in the United States to estimate the relationship between long-term exposure to PM^2.5^ and COVID-19 death rates. The results indicate that long-term exposure to air pollution increases vulnerability to the most severe COVID-19 outcomes. We found statistically significant evidence that an increase of 1 *μ* g/m^3^ in long-term PM^2.5^ exposure is associated with an 8% increase in the COVID-19 mortality rate. Our results were adjusted for a large set of socioeconomic, demographic, weather, behavioral, epidemic stage, social isolation measures, and healthcare-related confounders and demonstrated robustness across a wide range of sensitivity analyses.

In our previous study^20^ of 60 million Americans older than 65 years of age, we found that a 1 *μ*g/m^3^ in long-term PM^2.5^ exposure is associated with a 0.73% increase in the rate of all-cause mortality. Therefore, the same small increase in long-term exposure to PM^2.5^ led to an increase in the COVID-19 death rate of a magnitude 11 times that estimated for all-cause mortality.

Our results are consistent with previous findings that air pollution exposure increases severe outcomes during infectious disease outbreaks. Ciencewicki and Jaspers^19^ provide a review of the epidemiologic and experimental literature linking air pollution to infectious disease. During the 2003 outbreak of Severe Acute Respiratory Syndrome (SARS), a type of coronavirus closely related to COVID-19, Cui et al^41^ reported that locations in China with a moderate or high long-term air pollution index (API) had SARS case fatality rates 126% and 71% higher, respectively, than locations with low API. Long-term particulate matter exposure has been associated with hospitalizations for pneumonia in the well-controlled quasi-experimental conditions provided by the closing of the Utah Valley Steel Mill,^42^ and a link between long-term PM^2.5^ exposure and pneumonia and influenza deaths was reported in a well-validated cohort study.^28^ Several studies have reported associations between short-term PM^2.5^ exposure and poor infectious disease outcomes,^43 44^ including higher hospitalization rates or increased medical encounters for influenza, pneumonia, and acute lower respiratory infections. In these studies and in the literature on the association between air pollution and chronic disease outcomes, relationships with long-term pollution exposure tend to be stronger than relationships with short-term exposure,^20 45 46^ and the large effect estimate in our study is consistent with this trend.

Relationships have also been detected between pollution exposures and severe outcomes in the context of past pandemics. Studies found particulate matter exposure to be associated with the mortality during the H1N1 influenza pandemic in 2009.^47 48^ Recent studies have even used historic data to show a relationship between air pollution from coal burning and mortality in the 1918 Spanish influenza pandemic.^49 50^

Although our study design cannot provide insight into the mechanisms underlying the relationship between PM^2.5^ and COVID-19 mortality, prior studies have shed light on the potential biological mechanisms that may explain the relationship between air pollution and viral outcomes.^19^ PM^2.5^ exposure is known to be associated with many of the cardiovascular and respiratory comorbidities that dramatically increase the risk of death in COVID-19 patients. We hypothesize that the effects captured here are largely mediated by these comorbidities and pre-existing PM-related inflammation and cellular damage,^46 51^ as suggested by a recent commentary.^52^ Experimental studies^19 53-56^ also suggest that exposure to pollution can suppress early immune responses to the infection, leading to later increases in inflammation and worse prognosis, which may also explain our findings. Some studies^57-59^ have suggested that air pollution can also proliferate the transmission of infectious disease. If COVID-19 spread is indeed impacted by air pollution levels, which is not yet known, some of the effects detected in our study could be mediated by this factor as well.

This analysis provides a timely characterization of the relationship between historical exposure to air pollution and COVID-19 deaths in the United States. Research on how modifiable factors may exacerbate COVID-19 symptoms and increase mortality risk is essential to guide policies and behaviors to minimize fatality related to the outbreak. Our analysis relies on up-to-date population-level COVID-19 data and well-validated air pollution exposure measures.

Strengths of this analysis include adjusting for a wide range of potential confounders and a demonstrated robustness of results to different model choices. Moreover, the analyses rely exclusively on data and code that are publicly available. This provides a platform for the scientific community to continue updating and expanding these analyses as the pandemic evolves and data accumulate.

It is important to acknowledge that this study has limitations, mainly due to the fact that this is an ecological study with data available at the county level and that this is a cross-sectional study. High quality, nationwide individual-level COVID-19 outcome data are unavailable at this time and for the foreseeable future, thus necessitating the use of an ecologic study design for these analyses. Due to the potential for ecologic bias, our results should be interpreted in the context of this design and should not be used to make individual-level inferential statements. Also, unmeasured confounding bias is a threat to the validity of our conclusions. Unfortunately, in the midst of a pandemic it is not feasible to design a study and collect the data at the ideal level of spatial and temporal resolution to minimize all sources of bias. Yet, conditional on the data available, we have endeavored to adjust for confounding bias by all of the most important factors, including population density, time since the beginning of the outbreak, social isolation measures, behavior, weather, age structure, ethnicity, access to health care, and socio-economic factors. We also conducted 68 additional analyses to assess the robustness of the results to many modelling choices. Furthermore, we computed the E-value to demonstrate that the confounding effect of any unmeasured confounder would need to be much stronger than that of any of our observed confounders in order to explain away the relationship between PM^2.5^ exposure and COVID-19 mortality rate. The calculation of the E-value provided reassurance that the presence of a strong unmeasured confounder is unlikely; however, this possibility cannot be ruled out completely.

The inability to accurately quantify the number of COVID-19 cases due to limited testing capacity presents another potential limitation. We instead used total population size as the denominator for our mortality rates, and we additionally adjusted our models for numerous anticipated proxies of outbreak size, including time since first reported COVID-19 case, time since stay-at-home order was issued, and population density.

To conduct the most rigorous possible studies of air pollution and health using ecologic data, it is critical to utilize areal units that minimize within-area exposure variability and maximize between-area exposure variability.^60 61^ We anticipated that our use of counties satisfies this criterion, because counties generally represent meaningful boundaries between urban, suburban, and rural areas. These population density-related delineations also often correspond to steep gradients in air pollution levels, thus maximizing across-unit exposure variability while minimizing within-unit variability. We also note that the use of long-term county-level exposure data in our study likely led to some degree of exposure misclassification. However, previous literature has found that using sub-county scale PM^2.5^ exposure in studies of mortality tends to either have no impact or to increase the strength of the associations between PM^2.5^ and mortality from various causes.^62^

Because of the many limitations, this study also provides justification for expanded follow-up investigations as more and higher-quality COVID-19 data become available. Such studies would include validation of our findings with other data sources and study types, as well as studies of biological mechanisms, impacts of PM^2.5^ exposure timing, and relationships between PM^2.5^ and other COVID-19 outcomes such as hospitalization. The results of this study also underscore the importance of continuing to enforce existing air pollution regulations. Based on our results, we anticipate a failure to do so could potentially increase the long-term COVID-19 death toll and hospitalizations, as well as further burden our healthcare system with other PM^2.5^-related death and disease that would draw resources away from COVID-19 patients.

## Data Availability

The data and code are publicly available.

https://github.com/wxwx1993/PM_COVID

## Acknowledgments

The computations in this paper were run on (1) the Odyssey cluster supported by the FAS Division of Science, Research Computing Group at Harvard University; and (2) the Research Computing Environment supported by the Institute for Quantitative Social Science in the Faculty of Arts and Sciences at Harvard University. The authors would like to thank Lena Goodwin and Stacey Tobin for editorial assistance in the preparation of this manuscript.

## References

1. Onder G, Rezza G, Brusaferro S. Case-fatality rate and characteristics of patients dying in relation to COVID-19 in Italy. JAMA 2020 doi: 10.1001/jama.2020.4683 [published Online First: 2020/03/24]

2. Centers for Disease Control and Prevention Covid-19 Response Team. Severe outcomes among patients with Coronavirus Disease 2019 (COVID-19) - United States, February 12-March 16, 2020. MMWR Morb Mortal Wkly Rep 2020;69(12):343–46. doi: 10.15585/mmwr.mm6912e2 [published Online First: 2020/03/28]

3. Guan WJ, Ni ZY, Hu Y, et al. Clinical characteristics of coronavirus disease 2019 in China. N Engl J Med 2020 doi: 10.1056/NEJMoa2002032 [published Online First: 2020/02/29]

4. Wu JT, Leung K, Bushman M, et al. Estimating clinical severity of COVID-19 from the transmission dynamics in Wuhan, China. Nat Med 2020;26(4):506–10. doi: 10.1038/s41591-020-0822-7 [published Online First: 2020/04/15]

5. Centers for Disease Control and Prevention. National Vital Statistics System. Provisional Death Counts for Coronavirus Disease (COVID-19) Available from: https://www.cdc.gov/nchs/nvss/vsrr/COVID19/.

6. Garg S, Kim L, Whitaker M, et al. Hospitalization rates and characteristics of patients hospitalized with laboratory-confirmed Coronavirus Disease 2019 - COVID-NET, 14 States, March 1-30, 2020. MMWR Morb Mortal Wkly Rep 2020;69(15):458–64. doi: 10.15585/mmwr.mm6915e3 [published Online First: 2020/04/17]

7. Zhou F, Yu T, Du R, et al. Clinical course and risk factors for mortality of adult inpatients with COVID-19 in Wuhan, China: a retrospective cohort study. Lancet 2020;395(10229):1054–62. doi: 10.1016/S0140-6736(20)30566-3 [published Online First: 2020/03/15]

8. Chen L, Liu HG, Liu W, et al. [Analysis of clinical features of 29 patients with 2019 novel coronavirus pneumonia]. Zhonghua Jie He He Hu Xi Za Zhi 2020;43(0):E005. doi: 10.3760/cma.j.issn.1001-0939.2020.0005 [published Online First: 2020/02/07]

9. Huang C, Wang Y, Li X, et al. Clinical features of patients infected with 2019 novel coronavirus in Wuhan, China. Lancet 2020;395(10223):497–506. doi: 10.1016/S0140-6736(20)30183-5 [published Online First: 2020/01/28]

10. Mehta P, McAuley DF, Brown M, et al. COVID-19: consider cytokine storm syndromes and immunosuppression. Lancet 2020;395(10229):1033–34. doi: 10.1016/S0140-6736(20)30628-0 [published Online First: 2020/03/21]

11. Ruan Q, Yang K, Wang W, et al. Clinical predictors of mortality due to COVID-19 based on an analysis of data of 150 patients from Wuhan, China. Intensive Care Med 2020 doi: 10.1007/s00134-020-05991-x [published Online First: 2020/03/04]

12. Wang D, Hu B, Hu C, et al. Clinical characteristics of 138 hospitalized patients with 2019 novel coronavirus-infected pneumonia in Wuhan, China. JAMA 2020 doi: 10.1001/jama.2020.1585 [published Online First: 2020/02/08]

13. Xu Z, Shi L, Wang Y, et al. Pathological findings of COVID-19 associated with acute respiratory distress syndrome. Lancet Respir Med 2020;8(4):420–22. doi: 10.1016/S2213-2600(20)30076-X [published Online First: 2020/02/23]

14. Diamond M, Peniston Feliciano HL, Sanghavi D, et al. Acute Respiratory Distress Syndrome (ARDS) [Updated 2020 Jan 5]. StatPearls [Internet]. Treasure Island, FL: StatPearls Publishing 2020. Available from https://www.ncbi.nlm.nih.gov/books/NBK436002/.

15. Shi S, Qin M, Shen B, et al. Association of cardiac injury with mortality in hospitalized patients with COVID-19 in Wuhan, China. JAMA Cardiol 2020 doi: 10.1001/jamacardio.2020.0950 [published Online First: 2020/03/27]

16. Klok FA, Kruip M, van der Meer Njm, et al. Incidence of thrombotic complications in critically ill ICU patients with COVID-19. Thromb Res 2020 doi: 10.1016/j.thromres.2020.04.013 [published Online First: 2020/04/16]

17. Health Effects Institute. State of Global Air 2019 2019 Available from: www.stateofglobalair.org accessed April 23 2020.

18. Brook RD, Franklin B, Cascio W, et al. Air pollution and cardiovascular disease: a statement for healthcare professionals from the Expert Panel on Population and Prevention Science of the American Heart Association. Circulation 2004;109(21):2655–71. doi: 10.1161/01.CIR.0000128587.30041.C8 [published Online First: 2004/06/03]

19. Ciencewicki J, Jaspers I. Air pollution and respiratory viral infection. Inhal Toxicol 2007;19(14):1135–46. doi: 10.1080/08958370701665434 [published Online First: 2007/11/08]

20. Di Q, Wang Y, Zanobetti A, et al. Air pollution and mortality in the Medicare population. N Engl J Med 2017;376(26):2513–22. doi: 10.1056/NEJMoa1702747 [published Online First: 2017/06/29]

21. Dominici F, Peng RD, Bell ML, et al. Fine particulate air pollution and hospital admission for cardiovascular and respiratory diseases. JAMA 2006;295(10):1127–34. doi: 10.1001/jama.295.10.1127 [published Online First: 2006/03/09]

22. Puett RC, Hart JE, Yanosky JD, et al. Chronic fine and coarse particulate exposure, mortality, and coronary heart disease in the Nurses’ Health Study. Environ Health Perspect 2009;117(11):1697–701. doi: 10.1289/ehp.0900572 [published Online First: 2010/01/06]

23. Sram RJ, Binkova B, Dejmek J, et al. Ambient air pollution and pregnancy outcomes: a review of the literature. Environ Health Perspect 2005;113(4):375–82. doi: 10.1289/ehp.6362 [published Online First: 2005/04/07]

24. Wellenius GA, Burger MR, Coull BA, et al. Ambient air pollution and the risk of acute ischemic stroke. Arch Intern Med 2012;172(3):229–34. doi: 10.1001/archinternmed.2011.732 [published Online First: 2012/02/15]

25. Rhee J, Dominici F, Zanobetti A, et al. Impact of long-term exposures to ambient PM2.5 and ozone on ARDS risk for older adults in the United States. Chest 2019;156(1):71–79. doi: 10.1016/j.chest.2019.03.017 [published Online First: 2019/03/31]

26. United States Environmental Protection Agency. Integrated Science Assessment (ISA) for Particulate Matter (Final Report, 2019). EPA/600/R-19/188. Washington, DC: US EPA 2019.

27. Brook RD, Rajagopalan S, Pope CA, et al. Particulate matter air pollution and cardiovascular disease. Circulation 2010;121(21):2331–78. doi: doi:10.1161/CIR.0b013e3181dbece1

28. Pope CA, Burnett RT, Thurston GD, et al. Cardiovascular mortality and long-term exposure to particulate air pollution. Circulation 2004;109(1):71–77. doi: doi:10.1161/01.CIR.0000108927.80044.7F

29. Pope CA, Coleman N, Pond ZA, et al. Fine particulate air pollution and human mortality: 25+ years of cohort studies. Environ Res 2020;183:108924. doi: https://doi.org/10.1016/j.envres.2019.108924

30. Kioumourtzoglou MA, Schwartz JD, Weisskopf MG, et al. Long-term PM2.5 exposure and neurological hospital admissions in the Northeastern United States. Environ Health Perspect 2016;124(1):23–9. doi: 10.1289/ehp.1408973 [published Online First: 2015/05/16]

31. Zanobetti A, Dominici F, Wang Y, et al. A national case-crossover analysis of the short-term effect of PM2.5 on hospitalizations and mortality in subjects with diabetes and neurological disorders. Environ Health 2014;13(1):38. doi: 10.1186/1476-069X-13-38 [published Online First: 2014/06/03]

32. Dong E, Du H, Gardner L. An interactive web-based dashboard to track COVID-19 in real time. Lancet Infect Dis 2020 doi: 10.1016/S1473-3099(20)30120-1 [published Online First: 2020/02/23]

33. van Donkelaar A, Martin RV, Li C, et al. Regional estimates of chemical composition of fine particulate matter using a combined geoscience-statistical method with information from satellites, models, and monitors. Environ Sci Technol 2019;53(5):2595–611. doi: 10.1021/acs.est.8b06392 [published Online First: 2019/01/31]

34. Booth JG, Casella G, Friedl H, et al. Negative binomial loglinear mixed models. Stat Modelling 2003;3(3):179–91. doi: 10.1191/1471082X03st058oa

35. R Core Team. R: A language and environment for statistical computing. Vienna, Austria: R Foundation for Statistical Computing 2020. Available from: https://www.R-project.org/.

36. Zhang X, Mallick H, Tang Z, et al. Negative binomial mixed models for analyzing microbiome count data. BMC Bioinformatics 2017;18(1):4. doi: 10.1186/s12859-016-1441-7 [published Online First: 2017/01/05]

37. Bates D, Machler M, Bolker B, et al. Fitting linear mixed-effects models using lme4. J Stat Software 2015;67(1) doi: 10.18637/jss.v067.i01

38. Bolker B. glmer.nb Fitting Negative Binomial GLMMs Available from: https://www.rdocumentation.org/packages/lme4/versions/1.1-23/topics/glmer.nb.

39. VanderWeele TJ, Ding P. Sensitivity analysis in observational research: introducing the E-value. Ann Intern Med 2017;167(4):268–74. doi: 10.7326/M16-2607 [published Online First: 2017/07/12]

40. Yancy CW. COVID-19 and African Americans. JAMA 2020 doi: 10.1001/jama.2020.6548 [published Online First: 2020/04/16]

41. Cui Y, Zhang ZF, Froines J, et al. Air pollution and case fatality of SARS in the People’s Republic of China: an ecologic study. Environ Health 2003;2(1):15. doi: 10.1186/1476-069X-2-15 [published Online First: 2003/11/25]

42. Pope CA, 3rd. Respiratory disease associated with community air pollution and a steel mill, Utah Valley. Am J Public Health 1989;79(5):623–8. doi: 10.2105/ajph.79.5.623 [published Online First: 1989/05/01]

43. Croft DP, Zhang W, Lin S, et al. Associations between source-specific particulate matter and respiratory infections in New York state adults. Environ Sci Technol 2020;54(2):975–84. doi: 10.1021/acs.est.9b04295 [published Online First: 2019/11/23]

44. Horne BD, Joy EA, Hofmann MG, et al. Short-term elevation of fine particulate matter air pollution and acute lower respiratory infection. Am J Respir Crit Care Med 2018;198(6):759–66. doi: 10.1164/rccm.201709-1883OC [published Online First: 2018/04/14]

45. Di Q, Dai L, Wang Y, et al. Association of short-term exposure to air pollution with mortality in older adults. JAMA 2017;318(24):2446–56. doi: 10.1001/jama.2017.17923 [published Online First: 2017/12/28]

46. Tsai DH, Riediker M, Berchet A, et al. Effects of short- and long-term exposures to particulate matter on inflammatory marker levels in the general population. Environ Sci Pollut Res Int 2019;26(19):19697–704. doi: 10.1007/s11356-019-05194-y [published Online First: 2019/05/13]

47. Morales KF, Paget J, Spreeuwenberg P. Possible explanations for why some countries were harder hit by the pandemic influenza virus in 2009 - a global mortality impact modeling study. BMC Infect Dis 2017;17(1):642. doi: 10.1186/s12879-017-2730-0 [published Online First: 2017/09/28]

48. Xu Z, Hu W, Williams G, et al. Air pollution, temperature and pediatric influenza in Brisbane, Australia. Environ Int 2013;59:384–8. doi: 10.1016/j.envint.2013.06.022 [published Online First: 2013/08/06]

49. Clay K, Lewis J, Severnini E. Pollution, infectious disease, and mortality: evidence from the 1918 Spanish influenza pandemic. J Econ Hist 2018;78(4):1179–209. doi: 10.1017/S002205071800058X [published Online First: 2018/10/02]

50. Clay K, Lewis J, Severnini E. What explains cross-city variation in mortality during the 1918 influenza pandemic? Evidence from 438 U.S. cities. Econ Hum Biol 2019;35:42–50. doi: 10.1016/j.ehb.2019.03.010 [published Online First: 2019/05/10]

51. Pope CA, 3rd, Bhatnagar A, McCracken JP, et al. Exposure to fine particulate air pollution is associated with endothelial injury and systemic inflammation. Circ Res 2016;119(11):1204–14. doi: 10.1161/CIRCRESAHA.116.309279 [published Online First: 2016/10/27]

52. Conticini E, Frediani B, Caro D. Can atmospheric pollution be considered a co-factor in extremely high level of SARS-CoV-2 lethality in Northern Italy? Environ Pollut 2020:114465. doi: 10.1016/j.envpol.2020.114465 [published Online First: 2020/04/10]

53. Becker S, Soukup JM. Exposure to urban air particulates alters the macrophage-mediated inflammatory response to respiratory viral infection. J Toxicol Environ Health A 1999;57(7):445–57. doi: 10.1080/009841099157539 [published Online First: 1999/09/24]

54. Kaan PM, Hegele RG. Interaction between respiratory syncytial virus and particulate matter in guinea pig alveolar macrophages. Am J Respir Cell Mol Biol 2003;28(6):697–704. doi: 10.1165/rcmb.2002-0115OC [published Online First: 2003/05/23]

55. Lambert AL, Mangum JB, DeLorme MP, et al. Ultrafine carbon black particles enhance respiratory syncytial virus-induced airway reactivity, pulmonary inflammation, and chemokine expression. Toxicol Sci 2003;72(2):339–46. doi: 10.1093/toxsci/kfg032 [published Online First: 2003/03/26]

56. Lambert AL, Trasti FS, Mangum JB, et al. Effect of preexposure to ultrafine carbon black on respiratory syncytial virus infection in mice. Toxicol Sci 2003;72(2):331–8. doi: 10.1093/toxsci/kfg031 [published Online First: 2003/03/28]

57. Chen PS, Tsai FT, Lin CK, et al. Ambient influenza and avian influenza virus during dust storm days and background days. Environ Health Perspect 2010;118(9):1211–6. doi: 10.1289/ehp.0901782 [published Online First: 2010/05/04]

58. Peng L, Zhao X, Tao Y, et al. The effects of air pollution and meteorological factors on measles cases in Lanzhou, China. Environ Sci Pollut Res Int 2020;27(12):13524–33. doi: 10.1007/s11356-020-07903-4 [published Online First: 2020/02/08]

59. Ye Q, Fu JF, Mao JH, et al. Haze is a risk factor contributing to the rapid spread of respiratory syncytial virus in children. Environ Sci Pollut Res Int 2016;23(20):20178–85. doi: 10.1007/s11356-016-7228-6 [published Online First: 2016/07/22]

60. Elliott P, Savitz DA. Design issues in small-area studies of environment and health. Environ Health Perspect 2008;116(8):1098–104. doi: 10.1289/ehp.10817 [published Online First: 2008/08/19]

61. Kunzli N, Tager IB. The semi-individual study in air pollution epidemiology: a valid design as compared to ecologic studies. Environ Health Perspect 1997;105(10):1078–83. doi: 10.1289/ehp.105-1470382 [published Online First: 1997/12/24]

62. Crouse DL, Erickson AC, Christidis T, et al. Evaluating the sensitivity of PM2.5-mortality associations to the spatial and temporal scale of exposure assessment. Epidemiology 2020;31(2):168–76. doi: 10.1097/EDE.0000000000001136 [published Online First: 2019/11/07]

